# Complementary DNA-contact and domain-destabilizing mechanisms of *PAX9* and *MSX1* variants in non-syndromic tooth agenesis: an AlphaFold 3, ThermoMPNN, and AlphaGenome study

**DOI:** 10.64898/2026.11.17.26368325

**Authors:** Filipe Martins Vono, Valeria Gonzaga Santos, Andre Naufel de Toledo

**Author notes:** **Correspondence:** Filipe Martins Vono, Centro Universitário de Belo Horizonte (UniBH), Belo Horizonte, Minas Gerais, Brazil.

## Abstract

Missense variants in the DNA-binding domains of *PAX9* and *MSX1* cause non-syndromic tooth agenesis, yet most substitutions remain variants of uncertain significance. We investigated the structural mechanisms of 39 pathogenic missense variants in the PAX9 paired domain (n = 32) and MSX1 homeodomain (n = 7) alongside 77 non-coding/splice ClinVar variants. We modelled protein–DNA complexes with AlphaFold 3, estimated folding stability changes (ΔΔG) with ThermoMPNN, and compared pathogenic variants with gnomAD v4 domain variants. AlphaFold 3 confidence metrics and mutant interfaces were insensitive to point mutations. Instead, pathogenic *PAX9* variants acted through two complementary mechanisms relative to recurrent population controls: loss of direct DNA contact (< 4 Å; 50% vs 17%; p = 0.0025) or domain destabilization (median ΔΔG 1.17 vs 0.34 kcal/mol; p = 4.6×10⁻⁵). Leave-one-out cross-validation selected a stable rule (DNA contact or ΔΔG ≥ 1.5 kcal/mol; apparent sensitivity 84%, specificity 79%) concordant with published DNA-binding assays for five of eight *PAX9* mutants; canonical AlphaMissense (> 0.564) showed 8%–21% specificity across both domains. Mechanism did not predict missing teeth, whereas missense variants were milder than null alleles (p = 0.001). AlphaGenome matched SpliceAI and jointly prioritized seven intronic variants.

## 1. Introduction

Non-syndromic tooth agenesis—the congenital absence of one or more permanent teeth without systemic anomalies—is the most common developmental anomaly of the human dentition. Database surveys have catalogued approximately 198 causal variants across 15 genes, with 91.9% clustering in seven core odontogenic loci: *AXIN2*, *EDA*, *LRP6*, *MSX1*, *PAX9*, *WNT10A*, and *WNT10B* (1). Within this network, *PAX9* and *MSX1* encode the two principal sequence-specific transcription factors. Pathogenic *PAX9* variants preferentially cause molar oligodontia affecting second permanent molars (2), whereas *MSX1* variants typically affect second premolars and maxillary first premolars or third molars (3).

Early tooth morphogenesis depends on reciprocal signalling between the oral ectoderm and cranial neural crest-derived ectomesenchyme. *Pax9* and *Msx1* are co-expressed in condensing dental mesenchyme, where they maintain mesenchymal *Bmp4* expression required for the bud-to-cap transition; homozygous deletion of either *Msx1* (4) or *Pax9* (5) in mice arrests odontogenesis at the bud stage. In humans, heterozygous loss-of-function alleles act in a dosage-sensitive manner, and PAX9 and MSX1 physically synergize to activate the *Bmp4* and *Msx1* promoters (6). Notably, certain pathogenic substitutions—such as PAX9 G51S—preserve monomeric DNA binding while abolishing synergistic transactivation with MSX1, showing that missense alleles can disrupt odontogenesis through distinct molecular routes (6). Genotype–phenotype syntheses have compiled 61 *PAX9* variants across 178 patients (2) and homeodomain-enriched *MSX1* substitutions such as R202P, T180I, and L211R (3, 7, 8). However, direct biochemical assays exist for few alleles—most notably eight paired-domain PAX9 mutants characterized by Wang et al. (6)—while variants of uncertain significance (VUS) continue to accumulate in ClinVar (9).

Disease-causing missense variants in transcription factors classically act through two complementary mechanisms first formalized in p53: “DNA-contact” substitutions that eliminate base or backbone interactions without unfolding the domain, and “structural” substitutions that destabilize the protein core (10, 11). In the homologous PAX6 paired domain in aniridia, Alibés et al. (12) used FoldX (13) to explain 44 of 50 (88%) pathogenic missense variants through predicted losses of folding stability or DNA binding and specificity. High-throughput binding surveys subsequently confirmed widespread affinity and specificity perturbations across paired-domain and homeodomain families (14, 15). Yet no systematic structural analysis has evaluated *PAX9* and *MSX1* variants using biomolecular complex predictors such as AlphaFold 3 (16), deep-learning stability models such as ThermoMPNN (17), Genome Aggregation Database (gnomAD) v4 population controls (18), AlphaMissense (19), or regulatory sequence models such as AlphaGenome (20) alongside SpliceAI (21).

Here, we integrated AlphaFold 3 protein–DNA modelling, ThermoMPNN saturation stability estimation, gnomAD v4 controls, and AlphaGenome regulatory scoring across *PAX9* and *MSX1*. Specifically, we aimed to: (i) test whether pathogenic missense variants segregate into complementary DNA-contact and domain-destabilizing mechanisms relative to population controls; (ii) evaluate the cross-validated stability of a combined structural rule against published DNA-binding assays and clinical tooth-loss severity; (iii) benchmark AlphaGenome against SpliceAI on non-coding ClinVar variants; and (iv) perform exploratory structural prioritization of domain VUS.

## 2. Material and methods

### 2.1 Variant curation and population control definition

We retrieved single-gene variant records for human *PAX9* and *MSX1* from ClinVar via National Center for Biotechnology Information (NCBI) E-utilities (9) on 15 November 2026 and mapped substitutions to canonical isoforms (UniProt P55771 for PAX9; P28360 for MSX1). Missense variants were cross-referenced against curated family reports in Intarak et al. (2) for *PAX9* and Table 2 of Liang et al. (3) for *MSX1*. We queried GRCh38 combined exome and genome allele counts (AC) and allele frequencies (AF) in gnomAD v4 (`dataset: gnomad_r4`) via the Broad Institute GraphQL API (18) on 15 November 2026 and retrieved precomputed AlphaMissense scores from AlphaFold Database proteome files (19). An attempted Leiden Open Variation Database (LOVD) query failed owing to server IP blocking (HTTP 403) and was excluded.

We assigned consolidated missense variants to four non-overlapping classes: P for non-conflicting ClinVar Pathogenic/Likely pathogenic assertions; P-lit for curated tooth-agenesis family variants (2, 3) with gnomAD AF < 10⁻⁴ and no ClinVar conflict; B for ClinVar Benign/Likely benign variants or gnomAD AF ≥ 0.001; and U for ClinVar VUS, conflicting entries, or unclassified population variants. Excluding translation initiation (start_lost, Met1) alleles and one reference-incompatible report (“PAX9 Asn16Ile”, Val16 in P55771) yielded 568 *PAX9* and 793 *MSX1* full-length missense entries. Structural analyses targeted the DNA-binding domains: PAX9 paired-domain residues 6–130 (UniProt annotation 4–130, bounded at residue 6 by template coverage) and MSX1 homeodomain residues 172–231, excluding four extra-domain pathogenic candidates (PAX9 Y143C, A168G; MSX1 G122E, P247S).

The final pathogenic domain set comprised 39 missense variants: 32 in *PAX9* (16 P, 16 P-lit) and 7 in *MSX1* (2 P, 5 P-lit). Population controls comprised all domain gnomAD v4 missense variants lacking P or P-lit labels, analysed across all observed alleles (AC ≥ 1: *PAX9*, n = 124; *MSX1*, n = 59) and recurrent alleles (AC ≥ 2: *PAX9*, n = 48; *MSX1*, n = 29; primary calibration set). All 124 *PAX9* controls and 58 of 59 *MSX1* controls (all except Likely benign S210N, AC = 143) carried label U, overlapping the domain U catalogue (*PAX9*, n = 138; *MSX1*, n = 72).

### 2.2 Biomolecular complex modelling with AlphaFold 3

We predicted two wild-type and 39 pathogenic mutant protein–DNA complexes (41 jobs) on 15 November 2026 using AlphaFold Server (AlphaFold 3, default parameters, automatic random seed, five diffusion samples per job; non-commercial research terms) (16). PAX9 paired-domain constructs (residues 6–137) were modelled with the recognition DNA duplex from the human PAX6–DNA crystal structure (Protein Data Bank [PDB] 6PAX) (22), and MSX1 homeodomain constructs (residues 166–237) with the murine Msx-1–DNA duplex (PDB 1IG7) (23). From each top-ranked sample (ranking_score), we recorded global interface predicted template modelling score (ipTM), predicted template modelling score (pTM), protein–DNA chain-pair ipTM, minimum inter-chain predicted aligned error (PAE), per-residue predicted local distance difference test (pLDDT), and clash flags (has_clash).

Minimum heavy-atom Euclidean distance from each wild-type residue to either DNA strand defined direct contact (< 4.0 Å), proximal (4.0–8.0 Å), and distal (> 8.0 Å) classes, verified across all five wild-type diffusion samples. Mutant interface perturbation was quantified as the percentage change in heavy-atom protein–DNA pairs (< 4.0 Å) relative to wild type.

### 2.3 Protein folding stability estimation

We estimated folding free-energy changes upon substitution (ΔΔG, kcal/mol; positive values destabilizing, mutant minus wild-type folding free energy) using ThermoMPNN (commit 370f76e, default weights thermoMPNN_default.pt, PyTorch 2.5.1 on CPU) (17) on isolated wild-type AlphaFold 3 protein chains. Site-saturation mutagenesis spanned all construct positions (2,508 PAX9 and 1,368 MSX1 single-amino-acid substitutions) with verified coordinate alignment. Planned secondary checks with DDMut failed owing to web-server internal errors, including on its documentation example.

### 2.4 Statistical analysis and threshold stability evaluation

We compared categorical proportions with two-sided Fisher’s exact tests and evaluated unique DNA-contact site enrichment against the 33% paired-domain baseline using exact binomial tests. Continuous distributions were compared via Mann–Whitney *U*, area under the receiver operating characteristic (ROC) curve (AUC), and two-sided *p*-values (scipy.stats.mannwhitneyu, exact when tie-free; tie- and continuity-corrected asymptotic otherwise), alongside Spearman’s *ρ*.

To combine mechanisms in *PAX9* (32 pathogenic vs 48 recurrent gnomAD AC ≥ 2 controls), variants were classified as rule-positive if < 4.0 Å from DNA or if ΔΔG met or exceeded a cut-off *c*. Eighty-fold leave-one-out cross-validation (LOOCV) selected *c* across {0, 0.25, …, 3.0} kcal/mol on each 79-variant training split by maximizing Youden’s index (*J* = sensitivity + specificity − 1). Sensitivity grids varied contact (3.5, 4.0, 5.0 Å) and ΔΔG (0.5, 1.0, 1.5 kcal/mol) cut-offs. We calculated Wilson 95% confidence intervals (CIs) for sensitivity and specificity and both Simel log-scale (24) and 20,000-iteration stratified bootstrap 95% CIs for positive and negative likelihood ratios (LR+, LR−). Given only seven pathogenic *MSX1* variants, the *PAX9*-derived cut-off (1.5 kcal/mol) was transferred to *MSX1* descriptively without LOOCV. Domain U variants were stratified into ClinVar-only VUS absent from gnomAD, gnomAD singletons (AC = 1), and recurrent gnomAD controls (AC ≥ 2). Analyses used Python 3.12.10 (numpy 2.5.4, scipy 1.18.1, matplotlib 3.11.2).

### 2.5 Mechanistic and clinical severity comparisons

We compared rule-positive predictions against electrophoretic mobility shift assay (EMSA) data from Wang et al. (6) for eight pathogenic *PAX9* paired-domain mutants (verified against Table 1 and Figure 3B of PMC2706687; EMSA score ≤ 1 defined binding loss) to assess mechanistic concordance. Clinical severity was evaluated using missing permanent teeth (excluding third molars; one summary value per *PAX9* variant) from Intarak et al. (2), comparing missense mechanism classes against each other and against 25 null (frameshift/nonsense) *PAX9* variants.

**Table 1.**
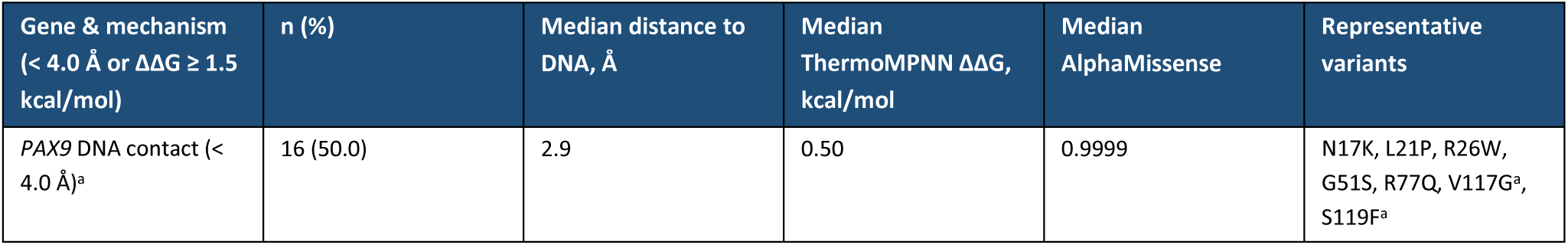
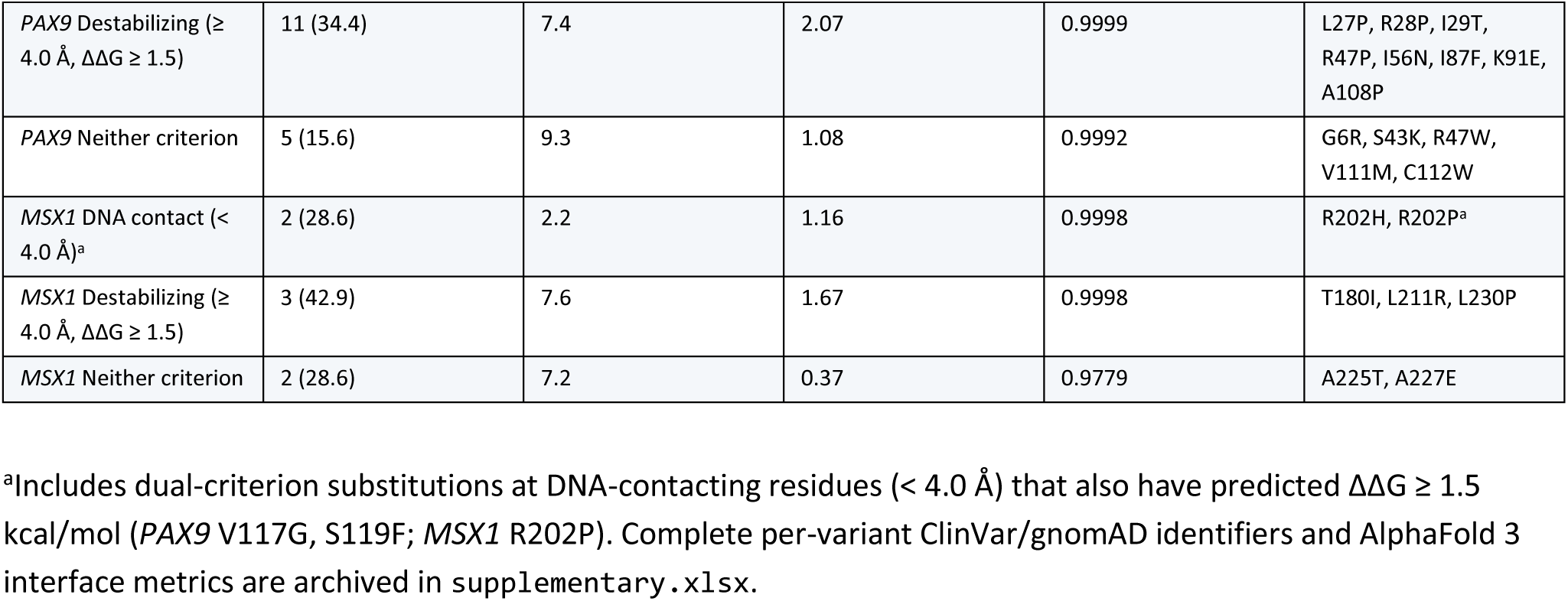
Mechanism-stratified summary of the 39 curated pathogenic missense variants in the PAX9 paired domain (residues 6–130, n = 32) and MSX1 homeodomain (residues 172–231, n = 7; full per-variant listing in Supplementary Table S1).

**Table 2.**
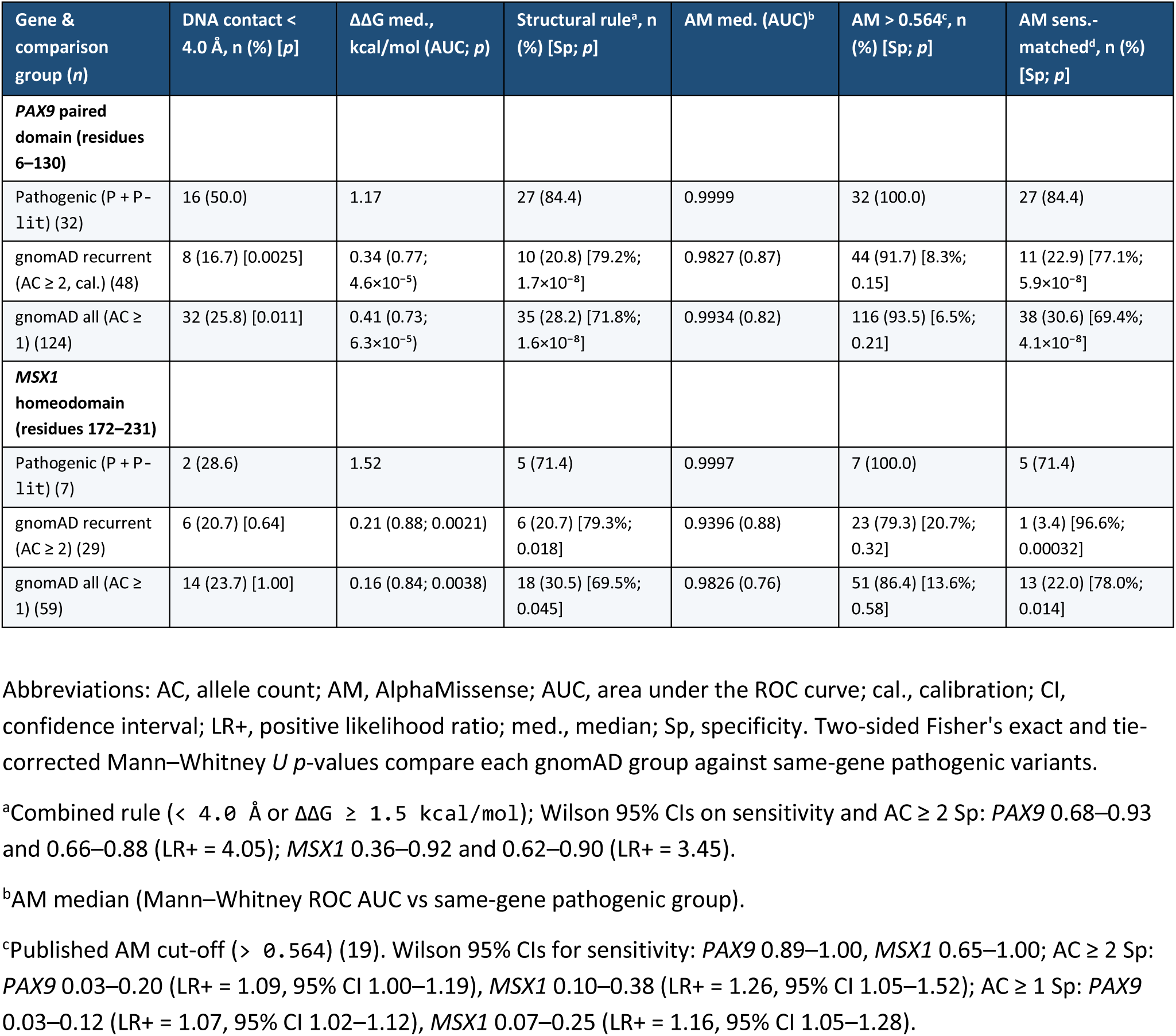
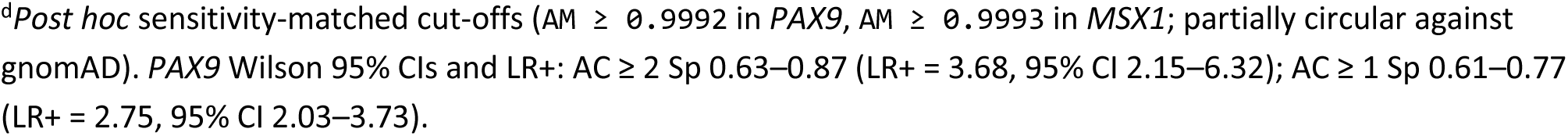
Comparison of curated pathogenic domain missense variants with gnomAD v4 population missense variants in the same domains.

### 2.6 Non-coding variant analysis with AlphaGenome and SpliceAI

From 84 single-gene non-coding or splice-region ClinVar variants with GRCh38 Sequence–Position–Deletion– Insertion (SPDI) coordinates, excluding three coding/boundary indels and three upstream intergenic *MSX1* entries left 78 variants with gene-restricted AlphaGenome 2 scores (7 Pathogenic/Likely pathogenic, 37 Benign/Likely benign, 34 VUS/unclassified [U]). Excluding translational start_lost *PAX9* g.36662092G>A (M1I) left 77 splicing variants (6 pathogenic, 37 benign, 34 U), of which 75 single-nucleotide variants had precomputed Ensembl Variant Effect Predictor SpliceAI Δ scores (21) (two small deletions lacked SpliceAI).

We scored variants within a 1 Mb window using AlphaGenome 2 (alphagenome v2.0.0, API v2, all folds; target-gene splice-site, splice-site usage, splice-junction, and RNA-seq scorers) and AlphaGenome 1 quantile scores (API v1) (20). Lacking odontogenic tracks, we benchmarked wild-type RNA-seq expression (99th percentile across 12 tracks) against *GAPDH* and *ALB* across mesenchymal stem cell (CL:0000134, primary condition), cranial neural crest cell (CL:0011012), osteoblast (UBERON:0002481_CL:0000062), and tongue (UBERON:0001723). AlphaFold Server and AlphaGenome outputs were generated strictly for non-commercial research and must not be used for clinical decisions.

### 2.7 Ethical considerations

This secondary computational study analysed only publicly accessible, non-identifiable aggregate and variant-level records from ClinVar, gnomAD v4, UniProt, PDB, and published summary tables (2, 3, 6). Because no human participants or biological specimens were involved, institutional ethics approval and informed consent were not required.

## 3. Results

### 3.1 Curated domain variant dataset

Variant curation identified 39 pathogenic missense substitutions within the analysed DNA-binding domains: 32 across 27 unique residues in the PAX9 paired domain (16 ClinVar P, 16 P-lit) and 7 across 6 unique residues in the MSX1 homeodomain (2 ClinVar P, 5 P-lit) (**Table 1**). Thirty-two of the 39 pathogenic alleles were absent from gnomAD v4, six were singletons (AC = 1), and one (*PAX9* K91E) had AC = 2. Domain control sets comprised 124 *PAX9* variants (48 recurrent with AC ≥ 2) and 59 *MSX1* variants (29 recurrent with AC ≥ 2).

### 3.2 Insensitivity of AlphaFold 3 mutant complexes to point substitutions

AlphaFold 3 generated high-confidence wild-type protein–DNA complexes, yet mutant complex models remained insensitive to pathogenic point substitutions (**Table 1**, **Supplementary Table S1**). Wild-type PAX9–DNA yielded ipTM = 0.86, pTM = 0.85, and 361 heavy-atom contacts across 44 interface residues (**Fig. 1A, 1C**), while wild-type MSX1–DNA yielded ipTM = 0.87, pTM = 0.88, and 179 contacts across 22 residues (**Fig. 1B**), with wild-type pLDDT values of 86.1–98.9 across all 39 mutated sites. However, re-predicting all 39 pathogenic mutants produced nearly unchanged global confidence scores (ipTM 0.84–0.87; pTM 0.83–0.89; |Δ protein–DNA chain-pair ipTM| ≤ 0.035; zero clashes across all 41 jobs) and modest interface contact changes (−5.8% to +9.1% in PAX9; −15.6% to +18.4% in MSX1). Critically, four *PAX9* substitutions that abolish DNA binding in vitro (6)—L21P (+9.1%), R26W (+0.3%), R28P (+0.6%), and I87F (+5.8%)—retained native-like predicted interfaces, motivating mechanistic analysis via wild-type DNA geometry and ThermoMPNN folding stability.

**Figure 1.**
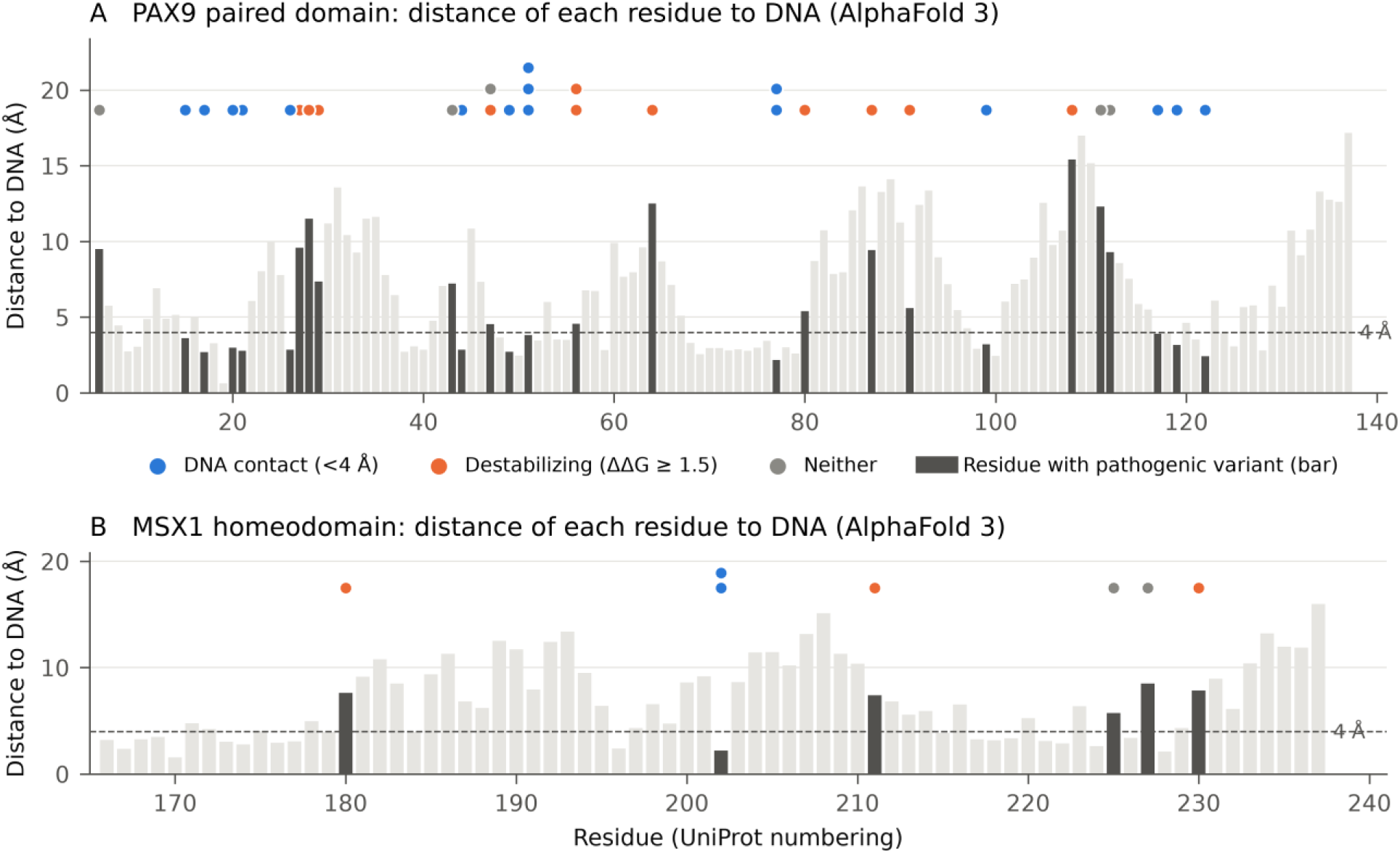
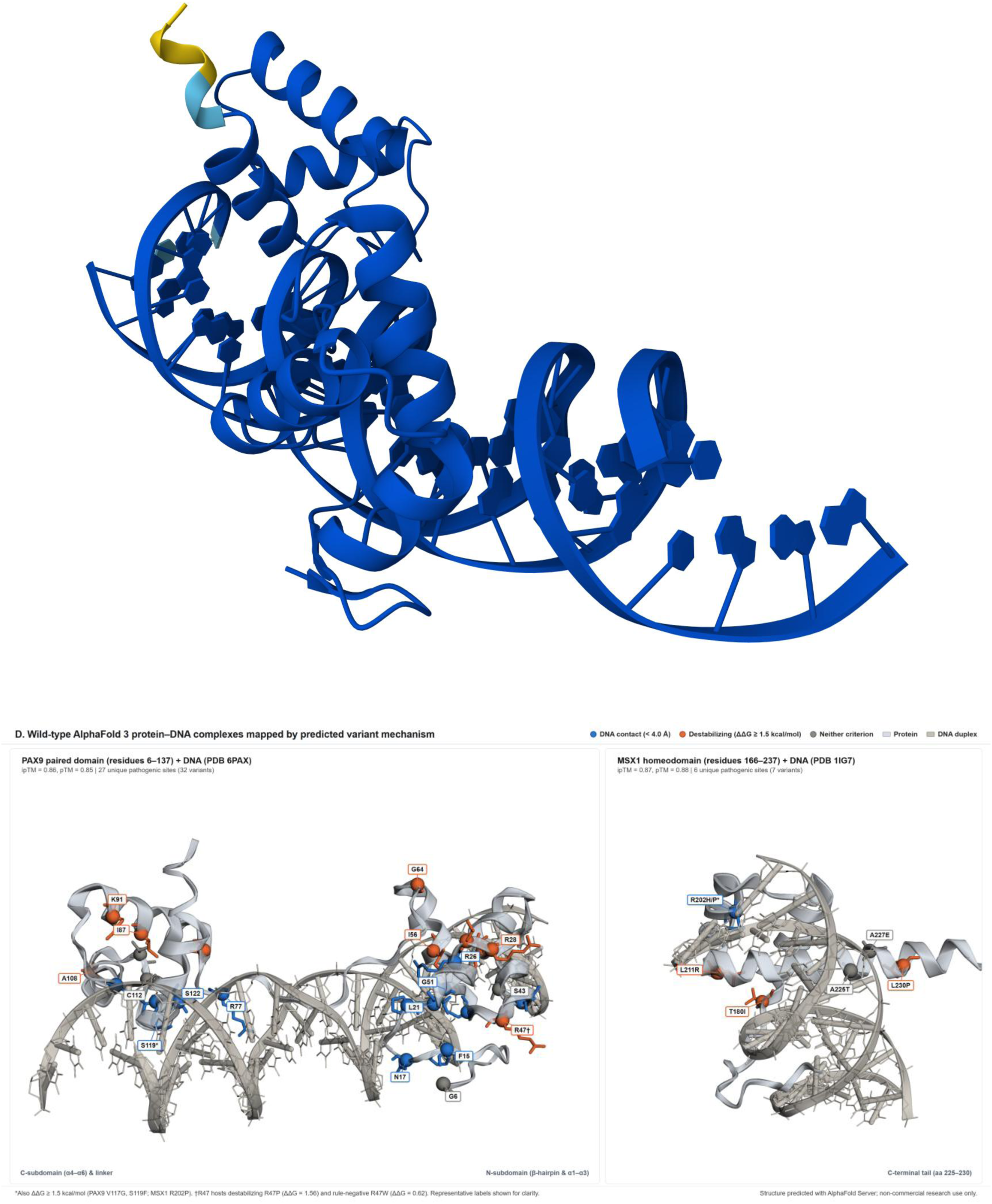
Residue-to-DNA distance profiles, AlphaFold 3 confidence, and three-dimensional mechanism mapping in PAX9 and MSX1. **(A, B)** Minimum heavy-atom distance from each wild-type residue to duplex DNA in top-ranked AlphaFold 3 complexes for the PAX9 paired domain (residues 6–137 with PDB 6PAX DNA) (22) **(A)** and MSX1 homeodomain (residues 166–237 with PDB 1IG7 DNA) (23) **(B)**. Darker bars mark residues with pathogenic variants, dashed lines mark the 4.0 Å contact cut-off, and circles classify variants as DNA contact (< 4.0 Å, blue), non-contact destabilizing (ΔΔG ≥ 1.5 kcal/mol, orange), or neither (grey). **(C)** Wild-type PAX9–DNA complex coloured by pLDDT (dark blue > 90, light blue 70–90, yellow 50–70; ipTM = 0.86, pTM = 0.85). **(D)** Three-dimensional mechanism mapping (3Dmol.js) across all 27 unique pathogenic PAX9 sites (left; 32 variants) and 6 MSX1 sites (right; 7 variants; ipTM = 0.87, pTM = 0.88); dual-criterion alleles (S119F*, V117G, R202P*) are blue and R47† (hosting destabilizing R47P and rule-negative R47W) is orange. *Structure predicted with AlphaFold Server; use is subject to the AlphaFold Server Output Terms of Use* (16).

**Figure 2.**
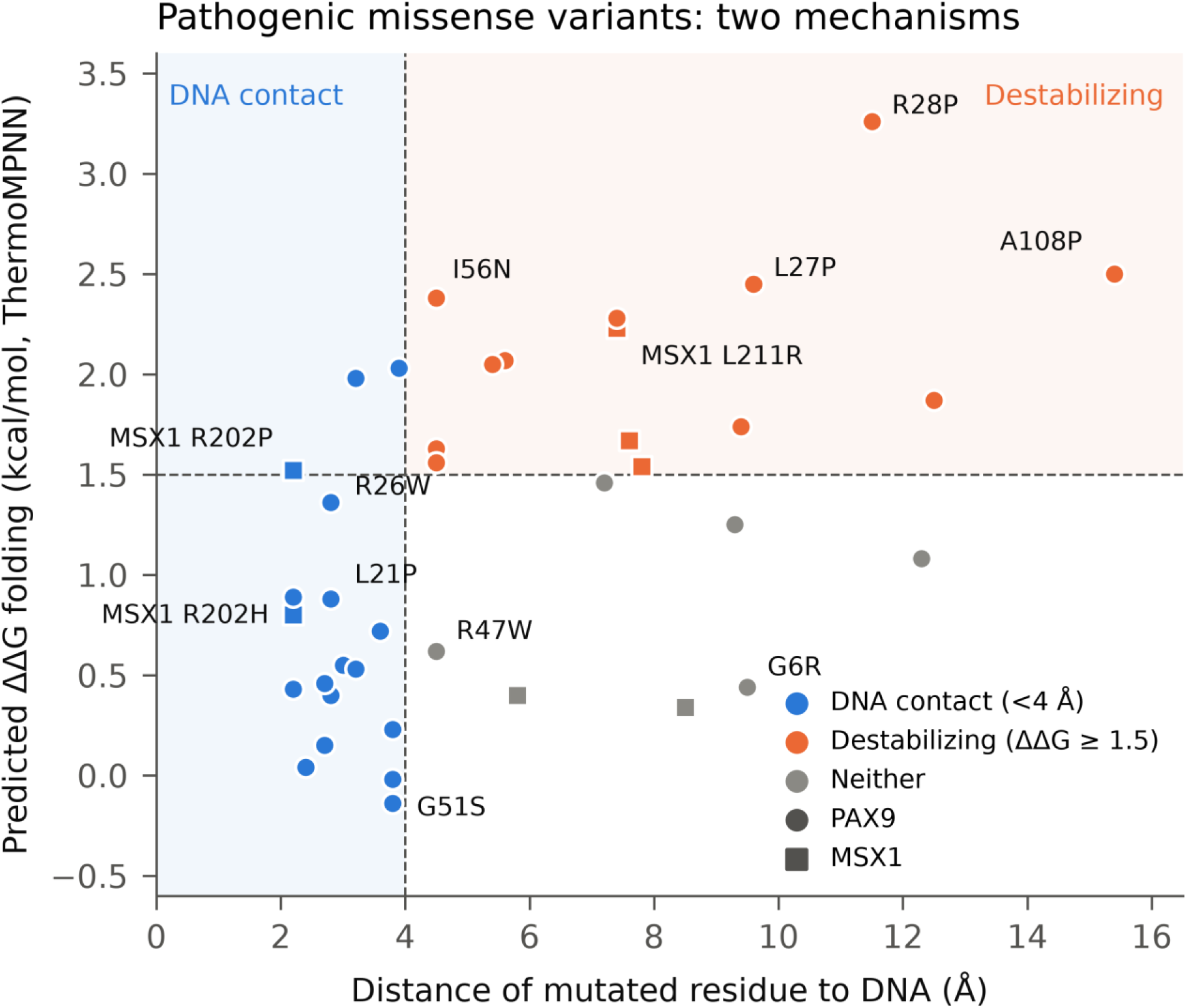
Complementary DNA-contact and domain-destabilizing mechanisms among pathogenic *PAX9* and *MSX1* missense variants. Wild-type minimum heavy-atom distance to DNA (Å) plotted against ThermoMPNN predicted folding stability change (ΔΔG, kcal/mol) (17) for curated pathogenic missense variants in the PAX9 paired domain (circles, n = 32) and MSX1 homeodomain (squares, n = 7). Dashed lines denote the 4.0 Å contact and cross-validated 1.5 kcal/mol destabilization thresholds. DNA-contacting *PAX9* variants show background-level ΔΔG (median 0.50 vs 0.56 kcal/mol across 2,508 substitutions), whereas non-contact variants are strongly destabilizing (median 1.81 kcal/mol; Mann–Whitney AUC = 0.88, *p* = 0.00028).

### 3.3 Two complementary mechanisms in the PAX9 paired domain and MSX1 homeodomain

Wild-type protein–DNA geometry partitioned pathogenic *PAX9* substitutions evenly between direct DNA-contact residues and interior scaffold positions (**Fig. 1A**, **Fig. 2**). Thirteen of the 27 unique pathogenic *PAX9* sites contacted DNA (< 4.0 Å; 48.1% vs 33.0% domain baseline; one-sided exact binomial *p* = 0.074; two-sided *p* = 0.103), with contact classification invariant across all five diffusion samples at 12 of 13 positions (varying only at Val117, 3.9 Å in the top-ranked sample). Across all 32 *PAX9* variants, 16 were direct DNA contacts (< 4.0 Å), 8 were proximal (4.0–8.0 Å), and 8 were distal (> 8.0 Å).

Integrating ThermoMPNN stability estimates revealed a strong inverse relationship between DNA proximity and domain destabilization (**Fig. 2**, **Table 1**). DNA-contacting *PAX9* variants preserved folding stability, showing a median ΔΔG of 0.50 kcal/mol (n = 16) matching the saturation-mutagenesis background across all 2,508 PAX9 substitutions (median 0.56 kcal/mol), exemplified by G51S (−0.14 kcal/mol) and N17K (0.15 kcal/mol). Conversely, non-contact *PAX9* variants were markedly destabilizing across both proximal (median 1.84 kcal/mol, n = 8) and distal sites (median 1.81 kcal/mol, n = 8; combined non-contact median 1.81 vs 0.50 kcal/mol; Mann–Whitney AUC = 0.88, *p* = 0.00028), led by helix-disrupting R28P (3.26 kcal/mol) and A108P (2.50 kcal/mol). Pathogenic *MSX1* homeodomain variants (n = 7) similarly showed elevated destabilization (median ΔΔG 1.52 vs 0.29 kcal/mol across 1,368 substitutions; AUC = 0.76), with four alleles in the top 16.2% of destabilizing changes—led by L211R (2.23 kcal/mol) and dual-criterion R202P (1.52 kcal/mol)—alongside DNA-contacting R202H (2.2 Å, 0.80 kcal/mol) and rule-negative A225T (0.40 kcal/mol, reported in autosomal recessive tooth agenesis) and A227E (0.34 kcal/mol) (**Fig. 1B**).

Projecting these mechanisms onto the three-dimensional AlphaFold 3 complexes confirmed clear spatial segregation between DNA-groove contacts and buried scaffold lesions (**Fig. 1D**). In PAX9, DNA-contact alleles traced the bipartite N-subdomain β-hairpin/helices, minor-groove linker, and C-subdomain recognition surface, whereas destabilizing alleles clustered within buried α-helical cores and rule-negative alleles occupied peripheral boundary or loop sites (**Fig. 1D, left**; **Table 1**). The MSX1 homeodomain displayed an analogous split between backbone-contacting R202 substitutions on helix α2, buried core lesions (T180I, L211R, L230P), and solvent-exposed α3 outer-face alleles (A225T, A227E) (**Fig. 1D, right**).

### 3.4 Comparison of pathogenic variants with gnomAD v4 population controls

Comparing pathogenic variants against recurrent gnomAD v4 domain controls (AC ≥ 2) demonstrated strong enrichment for both structural mechanisms (**Table 2**, **Fig. 3**). Pathogenic *PAX9* variants were three-fold more likely than recurrent controls to alter a DNA-contacting residue (50.0% [16/32] vs 16.7% [8/48]; Fisher’s exact *p* = 0.0025) and caused greater destabilization (median ΔΔG 1.17 vs 0.34 kcal/mol; AUC = 0.77, Mann–Whitney *p* = 4.6×10⁻⁵; ΔΔG ≥ 1.5 kcal/mol in 40.6% [13/32] vs 4.2% [2/48]; **Fig. 3A, 3B**). Under the primary combined rule (< 4.0 Å or ΔΔG ≥ 1.5 kcal/mol), 84.4% (27/32) of pathogenic *PAX9* variants tested positive versus 20.8% (10/48) of recurrent controls (Fisher’s exact *p* = 1.7×10⁻⁸; at ΔΔG ≥ 1.0 kcal/mol, 93.8% [30/32] vs 33.3% [16/48], *p* = 3.4×10⁻⁸), with consistent separation across all AC ≥ 1 controls (n = 124; **Table 2**). In *MSX1*, pathogenic variants were significantly more destabilizing than recurrent controls (median ΔΔG 1.52 vs 0.21 kcal/mol; AUC = 0.88, *p* = 0.0021), whereas DNA-contact proportions did not differ (28.6% [2/7] vs 20.7% [6/29]; *p* = 0.64).

**Figure 3.**
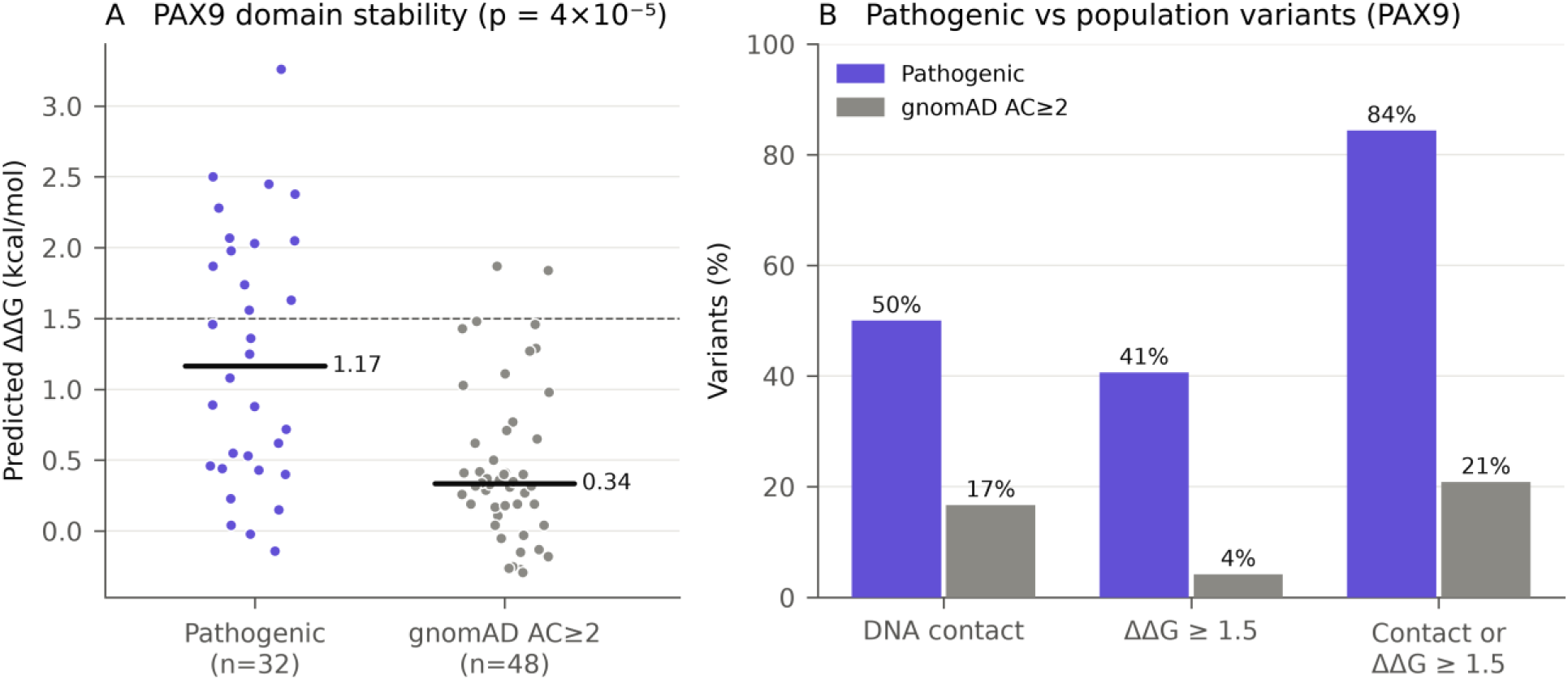
Enrichment of DNA-contact and destabilizing substitutions in pathogenic *PAX9* variants versus recurrent gnomAD v4 controls. **(A)** ThermoMPNN ΔΔG distributions for pathogenic *PAX9* paired-domain variants (n = 32) versus recurrent gnomAD v4 domain controls (AC ≥ 2, n = 48; horizontal bars mark medians: 1.17 vs 0.34 kcal/mol; Mann–Whitney *p* = 4.6×10⁻⁵). **(B)** Proportions of pathogenic *PAX9* variants versus recurrent controls at DNA-contact residues (< 4.0 Å; 50.0% vs 16.7%; Fisher’s exact *p* = 0.0025), with ΔΔG ≥ 1.5 kcal/mol (40.6% vs 4.2%), or positive under the combined structural rule (84.4% vs 20.8%; *p* = 1.7×10⁻⁸).

Although AlphaMissense achieved strong rank discrimination (AUC = 0.87 in *PAX9* and 0.88 in *MSX1* vs AC ≥ 2 controls), high sequence conservation saturated scores across neutral population controls (medians 0.983–0.993 in *PAX9*; 0.940–0.983 in *MSX1*; **Table 2**). At its canonical pathogenic threshold (> 0.564) (19), AlphaMissense classified 100.0% of pathogenic alleles (32/32 *PAX9*, 7/7 *MSX1*) as positive, but yielded specificities of only 8.3% on recurrent *PAX9* controls (44/48 positive, *p* = 0.15, LR+ = 1.09; AC ≥ 1 specificity 6.5% [116/124], *p* = 0.21, LR+ = 1.07) and 20.7% on recurrent *MSX1* controls (23/29 positive, *p* = 0.32, LR+ = 1.26; AC ≥ 1 specificity 13.6% [51/59], *p* = 0.58, LR+ = 1.16). Matching structural-rule sensitivity required extreme *post hoc* cut-offs of ≥ 0.9992 in *PAX9* (27/32 [84.4%]; specificity 77.1% [37/48], *p* = 5.9×10⁻⁸, LR+ = 3.68 on AC ≥ 2; 69.4% [86/124], *p* = 4.1×10⁻⁸, LR+ = 2.75 on AC ≥ 1) and ≥ 0.9993 in *MSX1* (5/7 [71.4%]; specificity 96.6% [28/29], *p* = 0.00032 on AC ≥ 2; 78.0% [46/59], *p* = 0.014 on AC ≥ 1), an optimistic recalibration because gnomAD frequencies inform AlphaMissense training (19).

### 3.5 Threshold stability in cross-validation and transfer to *MSX1*

Leave-one-out cross-validation confirmed complete threshold stability across all 80 training splits in *PAX9*. Maximizing Youden’s index selected ΔΔG ≥ 1.5 kcal/mol in every fold, yielding identical out-of-fold and apparent performance: sensitivity 0.84 (27/32; 95% Wilson CI 0.68–0.93), specificity 0.79 (38/48; 95% Wilson CI 0.66–0.88), LR+ 4.05 (Simel 95% CI 2.29–7.17; bootstrap 95% CI 2.50–8.40), and LR− 0.20 (Simel 95% CI 0.09–0.45; bootstrap 95% CI 0.04–0.38). Varying contact (3.5–5.0 Å) and ΔΔG (0.5–1.5 kcal/mol) cut-offs produced predictable trade-offs between sensitivity (0.72–0.97) and specificity (0.54–0.83) (**Supplementary Table S2**). Transferring the *PAX9*-derived 1.5 kcal/mol cut-off without re-fitting to *MSX1* (7 pathogenic vs 29 recurrent controls) yielded sensitivity 0.71 (5/7; 95% Wilson CI 0.36–0.92), specificity 0.79 (23/29; 95% Wilson CI 0.62–0.90), LR+ 3.45 (Simel 95% CI 1.47–8.10), and LR− 0.36 (Simel 95% CI 0.11–1.18).

### 3.6 Mechanistic concordance with published *PAX9* DNA-binding assays

Structural predictions matched published in vitro EMSA DNA-binding phenotypes for five of eight characterized *PAX9* paired-domain mutants (6) (**Table 3**). Four mutants with severe or complete binding loss tested positive via direct DNA contact (L21P, R26W) or core destabilization (R28P, I87F), whereas binding-competent G6R (three missing teeth; 9.5 Å, ΔΔG = 0.44 kcal/mol) was correctly rule-negative. Among the three discordant variants, S43K (7.2 Å, reduced EMSA binding +) had ΔΔG = 1.46 kcal/mol—0.04 kcal/mol below the 1.5 kcal/mol cut-off—and became concordant (6/8) at ΔΔG ≥ 1.0 kcal/mol. The remaining two exceptions acted downstream of monomeric DNA binding: G51S (3.8 Å, ΔΔG = −0.14 kcal/mol) retained EMSA binding while abolishing MSX1 synergy in a dominant-negative manner, and K91E (ΔΔG = 2.07 kcal/mol) retained EMSA binding while losing transactivation activity (6). Across all eight mutants, ThermoMPNN ΔΔG correlated moderately with stability changes modelled by Wang et al. (6) (*ρ* = 0.55), while experimental DNA binding correlated inversely with missing teeth (*ρ* = −0.72).

**Table 3.**
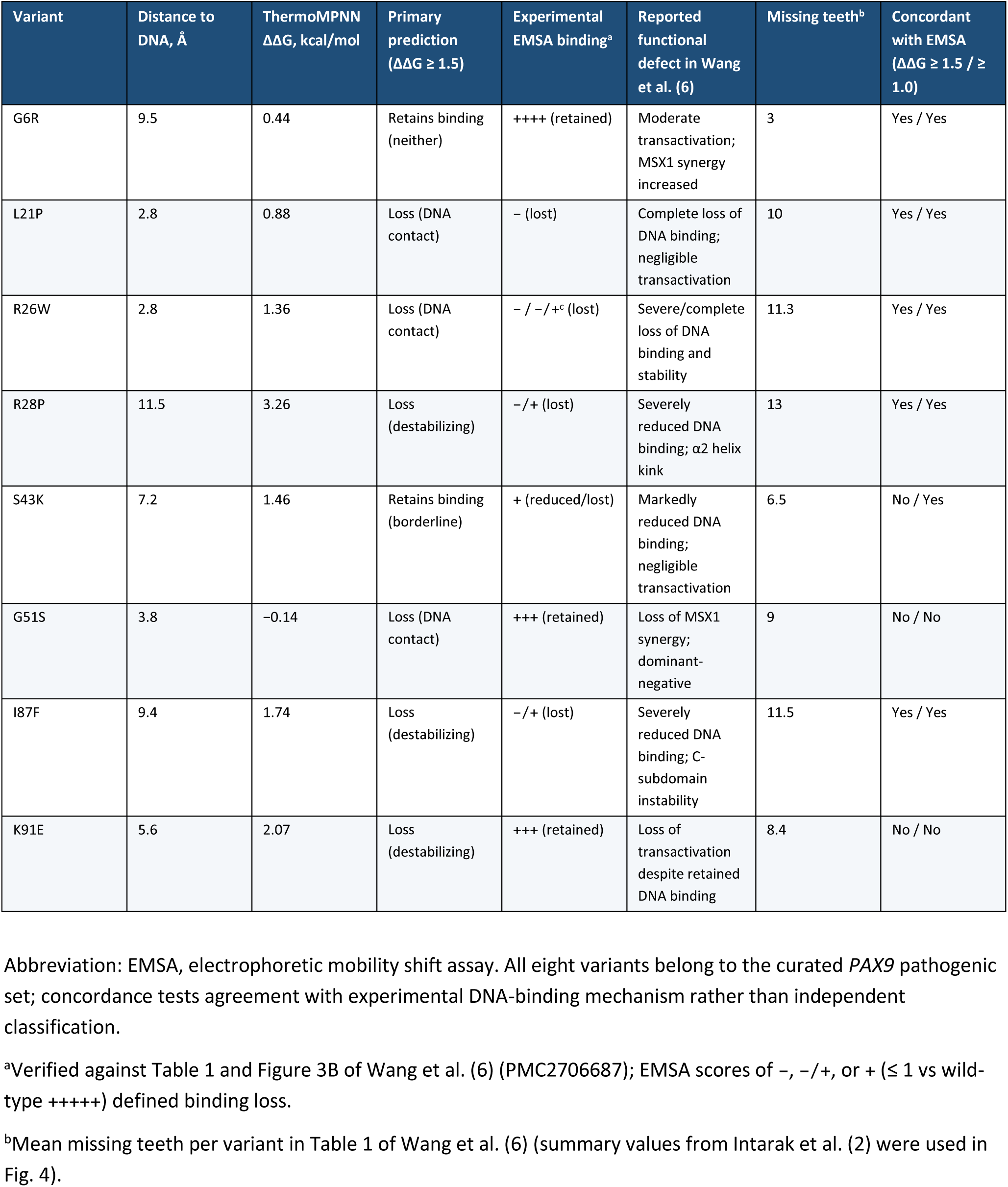
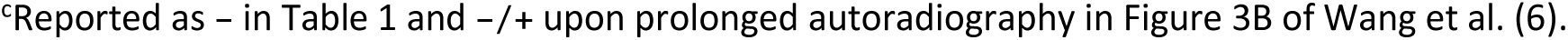
Mechanistic comparison of structural predictions with experimental functional assays for eight *PAX9*paired-domain mutants from Wang et al. (**6**).

| Variant | Distance to DNA, Å | ThermoMPNN ΔΔG, kcal/mol | Primary prediction (ΔΔG ≥ 1.5) | Experimental EMSA binding<sup>a</sup> | Reported functional defect in Wang et al. (6) | Missing teeth<sup>b</sup> | Concordant with EMSA (ΔΔG ≥ 1.5 / ≥ 1.0) |
|---|---|---|---|---|---|---|---|
| G6R | 9.5 | 0.44 | Retains binding (neither) | ++++ (retained) | Moderate transactivation; MSX1 synergy increased | 3 | Yes / Yes |
| L21P | 2.8 | 0.88 | Loss (DNA contact) | − (lost) | Complete loss of DNA binding; negligible transactivation | 10 | Yes / Yes |
| R26W | 2.8 | 1.36 | Loss (DNA contact) | − / −/+<sup>c</sup> (lost) | Severe/complete loss of DNA binding and stability | 11.3 | Yes / Yes |
| R28P | 11.5 | 3.26 | Loss (destabilizing) | −/+ (lost) | Severely reduced DNA binding; α2 helix kink | 13 | Yes / Yes |
| S43K | 7.2 | 1.46 | Retains binding (borderline) | + (reduced/lost) | Markedly reduced DNA binding; negligible transactivation | 6.5 | No / Yes |
| G51S | 3.8 | −0.14 | Loss (DNA contact) | +++ (retained) | Loss of MSX1 synergy; dominant-negative | 9 | No / No |
| I87F | 9.4 | 1.74 | Loss (destabilizing) | −/+ (lost) | Severely reduced DNA binding; C-subdomain instability | 11.5 | Yes / Yes |
| K91E | 5.6 | 2.07 | Loss (destabilizing) | +++ (retained) | Loss of transactivation despite retained DNA binding | 8.4 | No / No |

Abbreviation: EMSA, electrophoretic mobility shift assay. All eight variants belong to the curated *PAX9* pathogenic set; concordance tests agreement with experimental DNA-binding mechanism rather than independent classification.
<sup>a</sup>Verified against Table 1 and Figure 3B of Wang et al. (6) (PMC2706687); EMSA scores of −, −/+, or + (≤ 1 vs wild-type +++++) defined binding loss.
<sup>b</sup>Mean missing teeth per variant in Table 1 of Wang et al. (6) (summary values from Intarak et al. (2) were used in Fig. 4).
<sup>c</sup>Reported as − in Table 1 and −/+ upon prolonged autoradiography in Figure 3B of Wang et al. (6).

### 3.7 Predicted mechanism versus clinical tooth-loss severity

Structural mechanism did not predict permanent tooth-loss severity among *PAX9* missense variants, whereas null alleles caused significantly more severe oligodontia (**Fig. 4**). Across 25 pathogenic paired-domain missense variants compiled by Intarak et al. (2), median missing permanent teeth (excluding third molars) were 9.5 for DNA-contact (n = 10), 11.0 for destabilizing (n = 11), and 7.0 for rule-negative variants (n = 4; destabilizing vs contact AUC = 0.61, Mann–Whitney *p* = 0.42; at ΔΔG ≥ 1.0 kcal/mol, 10.0 vs 9.5, *p* = 0.60), and ΔΔG correlated weakly with tooth loss (*ρ* = 0.23). Conversely, frameshift and nonsense *PAX9* alleles (n = 25) caused more missing teeth than missense alleles (median 15.0 vs 10.0; AUC = 0.77, *p* = 0.0009 [displayed as *p* = 0.001 in Fig. 4]).

**Figure 4.**
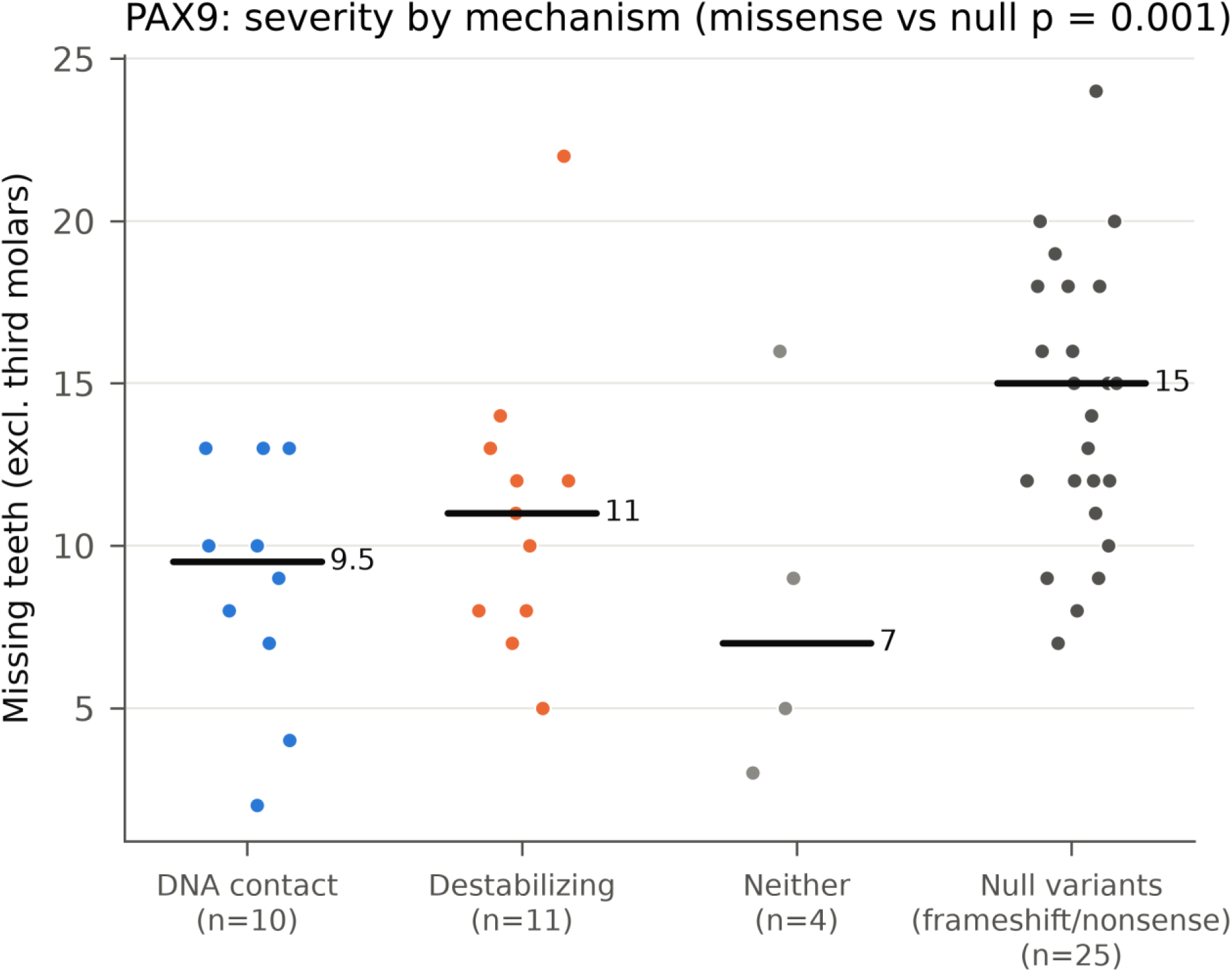
Clinical tooth-loss severity across predicted *PAX9* missense mechanisms and null alleles. Congenitally missing permanent teeth (excluding third molars; one summary value per variant from Intarak et al. (2)) across DNA-contact (< 4.0 Å, n = 10, median 9.5), non-contact destabilizing (ΔΔG ≥ 1.5 kcal/mol, n = 11, median 11.0), and rule-negative missense variants (n = 4, median 7.0) versus frameshift/nonsense null variants (n = 25, median 15.0; horizontal bars mark medians). Missense mechanism did not predict tooth loss (Mann–Whitney *p* = 0.42), whereas pooled missense alleles (n = 25, median 10.0) were milder than null alleles (AUC = 0.77, *p* = 0.0009; rounded to *p* = 0.001 in plot title).

### 3.8 Non-coding variant evaluation with AlphaGenome and SpliceAI

AlphaGenome matched SpliceAI on non-coding splice-variant discrimination, while wild-type expression benchmarking separated transcriptionally active from silent cell ontology tracks (**Fig. 5**, **Supplementary Table S3**). Mesenchymal stem cells (CL:0000134) expressed both *PAX9* (11.1× *ALB*) and *MSX1* (34.6× *ALB*), as did osteoblasts (UBERON:0002481_CL:0000062; *PAX9* 6.8×, *MSX1* 54.8× *ALB*), whereas cranial neural crest cells (CL:0011012) lacked *PAX9* expression (0.7× *ALB* vs 19.0× for *MSX1*) and tongue (UBERON:0001723) showed elevated *ALB* background (99th percentile 1.95). Across all 78 scored ClinVar variants, maximum AlphaGenome 2 splicing yielded AUC = 0.988. Excluding start_lost *PAX9* g.36662092G>A (leaving n = 77: 6 pathogenic, 37 benign, 34 U), AlphaGenome 2 splice-site, AlphaGenome 1 quantile, and SpliceAI Δ (n = 75 single-nucleotide variants: 5 pathogenic, 36 benign, 34 U) each achieved AUC = 1.000, while AlphaGenome 2 RNA-seq yielded AUC = 0.982 (0.94–0.99 across conditions).

**Figure 5.**
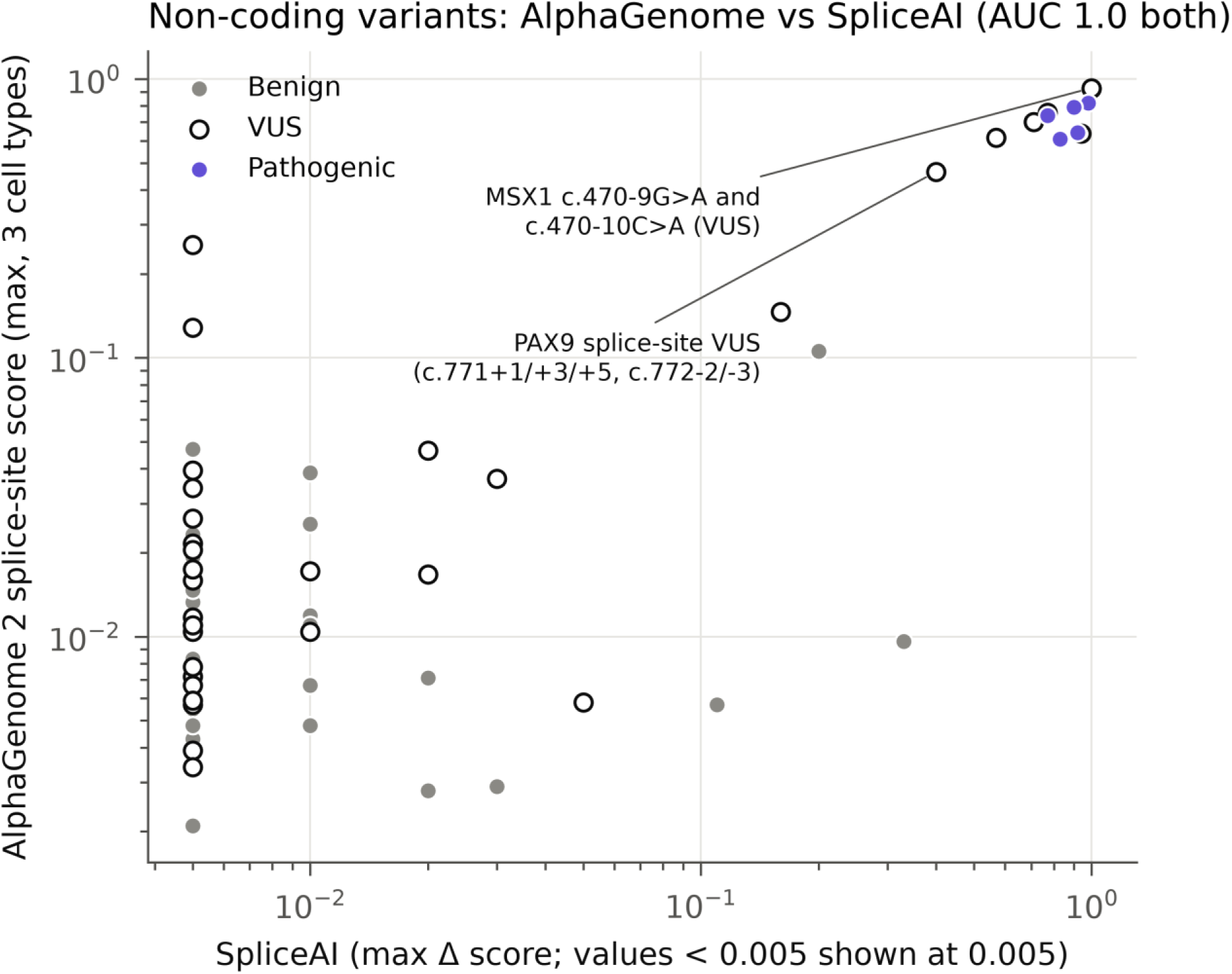
Concordance between AlphaGenome 2 and SpliceAI on non-coding *PAX9* and *MSX1* ClinVar variants. Log-scale comparison of maximum SpliceAI Δ score (< 0.005 plotted at 0.005) (21) against maximum AlphaGenome 2 gene-restricted splice-site score across mesenchymal stem cell, cranial neural crest cell, and tongue conditions (20) for single-nucleotide non-coding/splice ClinVar variants (n = 75: 5 pathogenic, violet; 36 benign, grey; 34 U, open circles; downstream prioritization required concordance across *PAX9*-expressing mesenchymal stem cell, osteoblast, and tongue tracks). Both predictors achieved AUC = 1.000 and jointly prioritized seven intronic U candidates (**Table 4**), including *MSX1* c.470-9G>A and c.470-10C>A.

**Table 4.** Exploratory, research-only prioritization of ClinVar-only domain missense variants (absent from gnomAD v4) and concordantly scored intronic/splice-region *PAX9* and *MSX1* candidates.^a^.

| Gene & variant | ClinVar ID (status; condition)<sup>b,c</sup> | DNA dist., Å / Splice region | ΔΔG, kcal/mol / AG2 (MSC / Ost / Tng) | AM / AG1 q | Primary prioritization (LR+ / SpliceAI Δ)<sup>d</sup> |
|---|---|---|---|---|---|
| **Part A. ClinVar-only domain missense variants absent from gnomAD v4 (AC = 0; rule-positive, n = 15)** |  |  |  |  |  |
| *PAX9* L32P | VCV003576569 (VUS<sup>b</sup>; Hypodontia, STHAG3) | 10.4 (distal) | 3.08 | 1.0000 | Destabilizing (LR+ 4.05) |
| *PAX9* G36V | VCV003654880 (VUS; Hypodontia) | 7.8 (proximal) | 1.82 | 0.9999 | Destabilizing (LR+ 4.05) |
| *PAX9* G73C | VCV000521952 (VUS; Inborn genetic diseases) | 2.9 (contact) | 0.55 | 0.9999 | DNA contact (LR+ 4.05) |
| *PAX9* V78L | VCV002002337 (VUS; Hypodontia) | 3.0 (contact) | −0.02 | 0.9994 | DNA contact (LR+ 4.05) |
| *PAX9* V83G | VCV001721820 (VUS; Hypodontia) | 7.8 (proximal) | 3.04 | 0.9992 | Destabilizing (LR+ 4.05) |
| *PAX9* V83L | VCV002575940 (VUS; Not provided) | 7.8 (proximal) | 1.53 | 0.9995 | Destabilizing (LR+ 4.05) |
| *PAX9* R88P | VCV004738820 (VUS; Hypodontia) | 13.3 (distal) | 1.85 | 0.9997 | Destabilizing (LR+ 4.05) |
| *PAX9* I97N | VCV001009882 (VUS; Hypodontia) | 4.3 (proximal) | 2.33 | 0.9993 | Destabilizing (LR+ 4.05) |
| *PAX9* R105P | VCV004685026 (VUS; Not provided) | 12.5 (distal) | 2.35 | 0.9999 | Destabilizing (LR+ 4.05) |
| *PAX9* L106R | VCV003777737 (VUS; STHAG3) | 9.8 (distal) | 3.19 | 0.9999 | Destabilizing (LR+ 4.05) |
| *MSX1* R176Q | VCV001971993 (VUS; EOHS) | 2.9 (contact) | 0.35 | 0.9996 | DNA contact (LR+ 3.45) |
| *MSX1* S206Y | VCV004530752 (VUS; Not provided) | 10.2 (distal) | 1.51 | 0.9992 | Destabilizing (LR+ 3.45) |
| *MSX1* K217R | VCV002631265 (VUS; MSX1-RD) | 3.3 (contact) | −0.12 | 0.9900 | DNA contact (LR+ 3.45) |
| *MSX1* W219R | VCV002078308 (Conflicting<sup>b</sup>; EOHS, MSX1-RD, STHAG1) | 3.4 (contact) | 2.61 | 1.0000 | DNA contact & Destab.<sup>d</sup> (LR+ 3.45) |
| *MSX1* K228E | VCV002574296 (VUS; Not provided) | 2.1 (contact) | 0.24 | 0.9998 | DNA contact (LR+ 3.45) |
| **Part B. Intronic/splice-region ClinVar group-U variants concordant across predictors (n = 7; columns show splice region, AG2 [MSC / Ost / Tng], AG1 q, and SpliceAI Δ)<sup>e</sup>** |  |  |  |  |  |
| *PAX9* c.771+1G>A | VCV004396677 (Unclassified<sup>e</sup>) | Splice donor (+1) | 0.087 / 0.170 / 0.638 | 1.0000 | SpliceAI Δ = 0.94 |
| *PAX9* c.771+3G>T | VCV002814643 (VUS) | Intron, donor region (+3) | 0.083 / 0.200 / 0.465 | 0.9995 | SpliceAI Δ = 0.40 |
| *PAX9* c.771+5G>C | VCV004921580 (VUS) | Intron, donor 5th base (+5) | 0.086 / 0.185 / 0.615 | 0.9996 | SpliceAI Δ = 0.57 |
| *PAX9* c.772-3C>G | VCV004396678 (Unclassified<sup>e</sup>) | Intron, polypyrimidine (−3) | 0.297 / 0.451 / 0.702 | 0.9999 | SpliceAI Δ = 0.71 |
| *PAX9* c.772-2A>G | VCV002821128 (VUS) | Splice acceptor (−2) | 0.354 / 0.589 / 0.757 | 1.0000 | SpliceAI Δ = 0.77 |
| *MSX1* c.470-10C>A | VCV004085302 (VUS) | Intron, cryptic acceptor (−10) | 0.924 / 0.957 / 0.888 | 1.0000 | SpliceAI Δ = 1.00 |
| *MSX1* c.470-9G>A | VCV002734641 (VUS) | Intron, cryptic acceptor (−9) | 0.928 / 0.958 / 0.889 | 1.0000 | SpliceAI Δ = 1.00 |

Abbreviations: AC, allele count in gnomAD v4 (gnomad\_r4); AG1 q, AlphaGenome 1 quantile score; AG2, AlphaGenome 2 gene-restricted splice-site score; AM, AlphaMissense score; Destab., destabilizing; dist., wild-type minimum heavy-atom distance to DNA; EOHS, hypoplastic enamel-onycholysis-hypohidrosis syndrome; LR+, nominal positive likelihood ratio; MSC, mesenchymal stem cell (CL:0000134); MSX1-RD, *MSX1*-related disorder; Ost, osteoblast (UBERON:0002481\_CL:0000062); STHAG1 / STHAG3, selective tooth agenesis 1 / 3; Tng, tongue (UBERON:0001723); VUS, variant of uncertain significance.
<sup>a</sup>Research-use-only disclaimer: outputs are exploratory, non-commercial research prioritizations under AlphaFold Server and AlphaGenome terms and must not be used for clinical diagnostic reporting. Part A lists all 15 rule-positive ClinVar-only U variants absent from gnomAD v4 (10/14 *PAX9*, 5/14 *MSX1*; all 210 U variants in Supplementary Table S4).
<sup>b</sup>Single-submitter VUS except multi-submitter non-conflicting *PAX9* L32P (VCV003576569) and conflicting *MSX1* W219R (VCV002078308).
<sup>c</sup>Submitter-reported ClinVar condition strings.
<sup>d</sup>*MSX1* W219R meets both criteria (3.4 Å and ΔΔG = 2.61 kcal/mol); nominal LR+ reflects *PAX9* calibration (4.05) and *MSX1* transfer (3.45).
<sup>e</sup>Transcripts NM\_001372076.1 (*PAX9*) and NM\_002448.3 (*MSX1*); VCV004396677 and VCV004396678 lacked a provided ClinVar significance string at retrieval (full 78-variant dataset in Supplementary Table S3).

All three splicing predictors concordantly prioritized seven intronic or splice-region group-U candidates (five VUS, two unclassified) across mesenchymal stem cell, osteoblast, and tongue tracks (AlphaGenome 1 quantile ≥ 0.9995; SpliceAI Δ 0.40–1.00; **Table 4**, **Supplementary Table S3**). These included proximal intronic *MSX1* variants NM_002448.3:c.470-9G>A and c.470-10C>A upstream of exon 2—predicted to create a strong cryptic acceptor (AlphaGenome 2 = 0.89–0.96 [0.93 and 0.92 in mesenchymal stem cells]; SpliceAI = 1.00)—alongside five *PAX9* variants flanking the c.771–c.772 intron junctions (led by NM_001372076.1:c.771+1G>A and c.772-2A>G). Three *PAX9* 3’ untranslated region VUS (c.*825T>G, c.*775C>T, c.*231T>G) exceeded background only in tongue (SpliceAI 0–0.16).

### 3.9 Exploratory structural characterization of domain VUS

Exploratory structural scoring of unclassified domain missense variants (U) revealed a steep enrichment gradient inversely tracking population allele frequency in *PAX9* (**Table 4**, **Supplementary Table S4**). Overall, 45 of 138 (32.6%) *PAX9* and 23 of 72 (31.9%) *MSX1* U variants were rule-positive. Stratified by population presence in *PAX9*, rule-positive proportions fell from 71.4% (10/14) among ClinVar-only VUS absent from gnomAD—exemplified by core-destabilizing L106R (ΔΔG = 3.19 kcal/mol) and L32P (3.08 kcal/mol) and DNA-contacting G73C (2.9 Å)—to 32.9% (25/76) among gnomAD singletons (AC = 1; e.g., R26Q at 2.8 Å, ΔΔG = 1.80 kcal/mol) and 20.8% (10/48) among recurrent AC ≥ 2 controls. Conversely, recurrent population alleles with saturated AlphaMissense scores— such as D94E (AC = 37, ΔΔG = 0.36 kcal/mol, AlphaMissense = 0.988) and E62K (AC = 10, ΔΔG = −0.25 kcal/mol, AlphaMissense = 0.9994)—tested cleanly rule-negative. In *MSX1*, rule-positive rates were 35.7% (5/14) among ClinVar-only candidates (including dual-criterion W219R [3.4 Å, ΔΔG = 2.61 kcal/mol] and DNA-contacting K228E [2.1 Å]; **Table 4**), 40.0% (12/30) among singletons (e.g., A227P, ΔΔG = 2.08 kcal/mol), and 21.4% (6/28) among recurrent U alleles (e.g., R224C, 2.6 Å).

## 4. Discussion

Pathogenic missense variants in the PAX9 paired domain—and preliminarily the MSX1 homeodomain—disrupt odontogenic transcription factor function through two largely mutually exclusive biophysical mechanisms: direct loss of DNA contacts or thermodynamic destabilization of the domain core. This dichotomy parallels classic contact versus structural p53 mutants (10, 11) and mirrors the FoldX analysis of homologous PAX6 in aniridia by

Alibés et al. (12, 13), and our cross-validated ThermoMPNN cut-off aligns closely with PAX6 energetic thresholds and ∼1–1.5 times ThermoMPNN’s benchmark root-mean-square error (17). Benchmarking against domain-matched human population controls in gnomAD v4 (18) proved critical, uncovering DNA-contact enrichment masked by uniform geometric baselines. Methodologically, AlphaFold 3 mutant complexes (16) retained native-like confidence and interface contacts even for biochemical null alleles, extending AlphaFold2’s known insensitivity to destabilizing point substitutions (25) to protein–DNA assemblies. Meanwhile, canonical AlphaMissense thresholds (19) suffered severe false-positive saturation across both conserved domains. Combining wild-type AlphaFold 3 DNA geometry with ThermoMPNN stability overcame both pitfalls without circular population-frequency recalibration, while distinguishing irreversible DNA-interface defects from scaffold-destabilizing mutants that retain an intact recognition surface and could theoretically respond to pharmacological chaperones (11, 12).

Both structural mechanisms produced similar missing-tooth counts among *PAX9* missense carriers, indicating that impaired DNA readout and core unfolding converge on reduced transcriptional output in dental mesenchyme (2, 12). By contrast, paired-domain missense variants caused milder oligodontia than frameshift or nonsense alleles, supporting dosage-sensitive residual hypomorphic activity in vivo (2, 6). Comparison with functional assays by Wang et al. (6) also clarified the boundaries of a two-feature monomeric model. Specifically, PAX9 G51S preserves monomeric DNA binding while abolishing synergistic co-activation of the *Bmp4* and *Msx1* promoters with MSX1 in a dominant-negative fashion, whereas K91E impairs transactivation downstream of DNA binding (6). Borderline substitutions near the stability threshold (such as S43K) further underscore the inherent uncertainty of hard cut-offs when ΔΔG prediction error is roughly 1 kcal/mol (17). Moreover, monomeric distance and stability metrics do not capture subtle shifts in DNA sequence specificity that preserve overall binding affinity across paired-domain and homeodomain proteins (12, 14, 15).

In non-coding regions of *PAX9* and *MSX1*, AlphaGenome (20) matched SpliceAI (21) on ClinVar splice-site classification without adding incremental discrimination over sequence-based splice modelling. Nevertheless, checking predicted baseline transcription prevented misinterpretation in cell tracks where *PAX9* was silent (cranial neural crest cells), and multi-tool concordance prioritized seven intronic candidates—notably cryptic-acceptor *MSX1* alleles upstream of exon 2 and five *PAX9* donor/acceptor variants—whereas evaluating distal cis-regulatory expression awaits odontogenic tissue tracks and confirmed regulatory variants. Under the Clinical Genome Resource (ClinGen) Bayesian framework for computational criteria PP3 and BP4 (26, 27), our nominal *PAX9* likelihood ratios approach supporting-to-moderate evidence strength, separating high-risk rare VUS from recurrent population variants where conservation models saturate. However, all structural and regulatory prioritizations reported here are strictly exploratory research outputs and must not be used for clinical diagnostic reporting.

Several statistical, modelling, and experimental limitations temper our conclusions and frame future validation. First, no independent external cohort of pathogenic and benign domain variants is currently available; the *MSX1* pathogenic sample is small (leaving its LR− confidence interval crossing unity); sample sizes yield wide confidence intervals; design choices (contact cut-off, disjunctive rule structure, and AC ≥ 2 calibration control definition) were established with dataset access; and overlap between recurrent gnomAD controls and domain U variants (all except Likely benign *MSX1* S210N) makes nominal likelihood ratios optimistic if applied back to calibration controls, whereas undiagnosed mild hypodontia in gnomAD makes case–control enrichment conservative. Second, ΔΔG estimates rely on a single tool (ThermoMPNN) (17) applied to apo protein chains, wild-type complexes use heterologous template DNA duplexes from PAX6 (22) and murine Msx-1 (23), minor gnomad_r4 subversion metadata were unlogged, single-submitter ClinVar labels may incorporate PP3 predictions, and AlphaGenome lacks primary tooth-germ tracks. Third, mechanistic and clinical comparisons relied on eight *PAX9* mutants within the calibration set (6) and heterogeneous literature tooth-count summaries (2) without new wet-laboratory experiments. Targeted quantitative EMSA, thermal-shift, MSX1-synergy reporter, and minigene splicing assays on the prioritized coding and intronic candidates (**Table 4**) are warranted to test these predictions experimentally.

## Data Availability

All Python analysis scripts, consolidated variant tables, ThermoMPNN saturation outputs, reduced AlphaFold Server archive bundles (verified byte-for-byte to reproduce all analytical tables), and processed AlphaGenome/SpliceAI score tables are openly archived on GitHub (https://github.com/contatofilipevono-rgb/agenesia-pax9-msx1). Structure predictions were generated with AlphaFold Server (use subject to non-commercial Output Terms of Use); AlphaGenome predictions were generated under non-commercial research terms. Outputs are for research only and must not be used for clinical diagnosis or patient management.

## Declarations

### Ethics statement

No DNA or biological samples were collected, no human participants were recruited, and no individual-level genotypes or identifiable clinical records were accessed. Only publicly accessible, fully anonymous, variant-level and aggregate data from public repositories and published literature were analysed; institutional ethics committee approval and informed consent were therefore not required.

### Data and code availability

All Python analysis scripts, consolidated variant tables, ThermoMPNN saturation outputs, reduced AlphaFold Server archive bundles (verified byte-for-byte to reproduce all analytical tables), and processed AlphaGenome/SpliceAI score tables are openly archived on GitHub (https://github.com/contatofilipevono-rgb/agenesia-pax9-msx1). Structure predictions were generated with AlphaFold Server: *“Structure predicted with AlphaFold Server; use is subject to the AlphaFold Server Output Terms of Use”* (non-commercial use only). AlphaGenome predictions were generated via the AlphaGenome API under non-commercial research terms of use; API keys are not distributed. All structural classifications, likelihood ratios, regulatory scores, and VUS prioritizations reported in this study are strictly for non-commercial research and hypothesis generation and must not be used for clinical variant reporting, genetic counseling, diagnostic interpretation, or patient management without independent experimental and clinical validation.

### Author contributions (CRediT)

**Filipe Martins Vono:** Conceptualization; Data curation; Formal analysis; Investigation; Methodology; Software; Visualization; Writing – original draft; Writing – review & editing. **Valeria Gonzaga Santos:** Validation; Writing – review & editing. **Andre Naufel de Toledo:** Validation; Writing – review & editing.

## References

1. Yu M, Wong SW, Han D, Cai T. Genetic analysis: Wnt and other pathways in nonsyndromic tooth agenesis. Oral Dis. 2019;25(3):646–651. doi:10.1111/odi.12931

2. Intarak N, Tongchairati K, Termteerapornpimol K, Chantarangsu S, Porntaveetus T. Tooth agenesis patterns and variants in PAX9: A systematic review. Jpn Dent Sci Rev. 2023;59:129–137. doi:10.1016/j.jdsr.2023.04.001

3. Liang J, Von den Hoff J, Lange J, Ren Y, Bian Z, Carels CE. MSX1 mutations and associated disease phenotypes: genotype-phenotype relations. Eur J Hum Genet. 2016;24(12):1663–1670. doi:10.1038/ejhg.2016.78

4. Satokata I, Maas R. Msx1 deficient mice exhibit cleft palate and abnormalities of craniofacial and tooth development. Nat Genet. 1994;6(4):348–356. doi:10.1038/ng0494-348

5. Peters H, Neubüser A, Kratochwil K, Balling R. Pax9-deficient mice lack pharyngeal pouch derivatives and teeth and exhibit craniofacial and limb abnormalities. Genes Dev. 1998;12(17):2735–2747. doi:10.1101/gad.12.17.2735

6. Wang Y, Groppe JC, Wu J, Ogawa T, Mues G, D’Souza RN, Kapadia H. Pathogenic mechanisms of tooth agenesis linked to paired domain mutations in human PAX9. Hum Mol Genet. 2009;18(15):2863–2874. doi:10.1093/hmg/ddp221

7. Vastardis H, Karimbux N, Guthua SW, Seidman JG, Seidman CE. A human MSX1 homeodomain missense mutation causes selective tooth agenesis. Nat Genet. 1996;13(4):417–421. doi:10.1038/ng0896-417

8. Yamaguchi S, Machida J, Kamamoto M, Kimura M, Shibata A, Tatematsu T, Miyachi H, Higashi Y, Jezewski P, Nakayama A, Shimozato K, Tokita Y. Characterization of novel MSX1 mutations identified in Japanese patients with nonsyndromic tooth agenesis. PLoS One. 2014;9(8):e102944. doi:10.1371/journal.pone.0102944

9. Landrum MJ, Chitipiralla S, Brown GR, Chen C, Gu B, Hart J, Hoffman D, Jang W, Kaur K, Liu C, Lyoshin V, Maddipatla Z, Maiti R, Mitchell J, O’Leary N, Riley GR, Shi W, Zhou G, Schneider V, Maglott D, Holmes JB, Kattman BL. ClinVar: improvements to accessing data. Nucleic Acids Res. 2020;48(D1):D835–D844. doi:10.1093/nar/gkz972

10. Cho Y, Gorina S, Jeffrey PD, Pavletich NP. Crystal structure of a p53 tumor suppressor-DNA complex: understanding tumorigenic mutations. Science. 1994;265(5170):346–355. doi:10.1126/science.8023157

11. Bullock AN, Fersht AR. Rescuing the function of mutant p53. Nat Rev Cancer. 2001;1(1):68–76. doi:10.1038/35094077

12. Alibés A, Nadra AD, De Masi F, Bulyk ML, Serrano L, Stricher F. Using protein design algorithms to understand the molecular basis of disease caused by protein-DNA interactions: the Pax6 example. Nucleic Acids Res. 2010;38(21):7422–7431. doi:10.1093/nar/gkq683

13. Schymkowitz JW, Rousseau F, Martins IC, Ferkinghoff-Borg J, Stricher F, Serrano L. Prediction of water and metal binding sites and their affinities by using the Fold-X force field. Proc Natl Acad Sci U S A. 2005;102(29):10147–10152. doi:10.1073/pnas.0501980102

14. Barrera LA, Vedenko A, Kurland JV, Rogers JM, Gisselbrecht SS, Rossin EJ, Woodard J, Mariani L, Kock KH, Inukai S, Siggers T, Shokri L, Gordân R, Sahni N, Cotsapas C, Hao T, Yi S, Kellis M, Daly MJ, Vidal M, Hill DE, Bulyk ML. Survey of variation in human transcription factors reveals prevalent DNA binding changes. Science. 2016;351(6280):1450–1454. doi:10.1126/science.aad2257

15. Kock KH, Kimes PK, Gisselbrecht SS, Inukai S, Phanor SK, Anderson JT, Ramakrishnan G, Lipper CH, Song D, Kurland JV, Rogers JM, Jeong R, Blacklow SC, Irizarry RA, Bulyk ML. DNA binding analysis of rare variants in homeodomains reveals homeodomain specificity-determining residues. Nat Commun. 2024;15(1):3110. doi:10.1038/s41467-024-47396-0

16. Abramson J, Adler J, Dunger J, Evans R, Green T, Pritzel A, Ronneberger O, Willmore L, Ballard AJ, Bambrick J, Bodenstein SW, Evans DA, Hung CC, O’Neill M, Reiman D, Tunyasuvunakool K, Wu Z, Žemgulytė A, Arvaniti E, Beattie C, Bertolli O, Bridgland A, Cherepanov A, Congreve M, Cowen-Rivers AI, Cowie A, Figurnov M, Fuchs FB, Gladman H, Jain R, Khan YA, Low CMR, Perlin K, Potapenko A, Savy P, Singh S, Stecula A, Thillaisundaram A, Tong C, Yakneen S, Zhong ED, Zielinski M, Žídek A, Bapst V, Kohli P, Jaderberg M, Hassabis D, Jumper JM. Accurate structure prediction of biomolecular interactions with AlphaFold 3. Nature. 2024;630(8016):493–500. doi:10.1038/s41586-024-07487-w

17. 17. Dieckhaus H, Brocidiacono M, Randolph NZ, Kuhlman B. Transfer learning to leverage larger datasets for improved prediction of protein stability changes. Proc Natl Acad Sci U S A. 2024;121(6):e2314853121. doi:10.1073/pnas.2314853121

18. Chen S, Francioli LC, Goodrich JK, Collins RL, Kanai M, Wang Q, Alföldi J, Watts NA, Vittal C, Gauthier LD, Poterba T, Wilson MW, Tarasova Y, Phu W, Grant R, Yohannes MT, Koenig Z, Farjoun Y, Banks E, Donnelly S, Gabriel S, Gupta N, Ferriera S, Tolonen C, Novod S, Bergelson L, Roazen D, Ruano-Rubio V, Covarrubias M, Llanwarne C, Petrillo N, Wade G, Jeandet T, Munshi R, Tibbetts K, Genome Aggregation Database Consortium, O’Donnell-Luria A, Solomonson M, Seed C, Martin AR, Talkowski ME, Rehm HL, Daly MJ, Tiao G, Neale BM, MacArthur DG, Karczewski KJ. A genomic mutational constraint map using variation in 76,156 human genomes. Nature. 2024;625(7993):92–100. doi:10.1038/s41586-023-06045-0

19. Cheng J, Novati G, Pan J, Bycroft C, Žemgulytė A, Applebaum T, Pritzel A, Wong LH, Zielinski M, Sargeant T, Schneider RG, Senior AW, Jumper J, Hassabis D, Kohli P, Avsec Ž. Accurate proteome-wide missense variant effect prediction with AlphaMissense. Science. 2023;381(6664):eadg7492. doi:10.1126/science.adg7492

20. Avsec Ž, Latysheva N, Cheng J, Novati G, Taylor KR, Ward T, Bycroft C, Nicolaisen L, Arvaniti E, Pan J, Thomas R, Dutordoir V, Perino M, De S, Karollus A, Gayoso A, Sargeant T, Mottram A, Wong LH, Drotár P, Kosiorek A, Senior A, Tanburn R, Applebaum T, Basu S, Hassabis D, Kohli P. Advancing regulatory variant effect prediction with AlphaGenome. Nature. 2026;649(8099):1206–1218. doi:10.1038/s41586-025-10014-0

21. Jaganathan K, Kyriazopoulou Panagiotopoulou S, McRae JF, Darbandi SF, Knowles D, Li YI, Kosmicki JA, Arbelaez J, Cui W, Schwartz GB, Chow ED, Kanterakis E, Gao H, Kia A, Batzoglou S, Sanders SJ, Farh KK. Predicting splicing from primary sequence with deep learning. Cell. 2019;176(3):535–548.e24. doi:10.1016/j.cell.2018.12.015

22. Xu HE, Rould MA, Xu W, Epstein JA, Maas RL, Pabo CO. Crystal structure of the human Pax6 paired domain-DNA complex reveals specific roles for the linker region and carboxy-terminal subdomain in DNA binding. Genes Dev. 1999;13(10):1263–1275. doi:10.1101/gad.13.10.1263

23. Hovde S, Abate-Shen C, Geiger JH. Crystal structure of the Msx-1 homeodomain/DNA complex. Biochemistry. 2001;40(40):12013– 12021. doi:10.1021/bi0108148

24. Simel DL, Samsa GP, Matchar DB. Likelihood ratios with confidence: sample size estimation for diagnostic test studies. J Clin Epidemiol. 1991;44(8):763–770. doi:10.1016/0895-4356(91)90128-v

25. Pak MA, Markhieva KA, Novikova MS, Petrov DS, Vorobyev IS, Maksimova ES, Kondrashov FA, Ivankov DN. Using AlphaFold to predict the impact of single mutations on protein stability and function. PLoS One. 2023;18(3):e0282689. doi:10.1371/journal.pone.0282689

26. Pejaver V, Byrne AB, Feng BJ, Pagel KA, Mooney SD, Karchin R, O’Donnell-Luria A, Harrison SM, Tavtigian SV, Greenblatt MS, Biesecker LG, Radivojac P, Brenner SE, ClinGen Sequence Variant Interpretation Working Group. Calibration of computational tools for missense variant pathogenicity classification and ClinGen recommendations for PP3/BP4 criteria. Am J Hum Genet. 2022;109(12):2163–2177. doi:10.1016/j.ajhg.2022.10.013

27. Ramadane-Morchadi L, Rotenberg N, Esteban-Sánchez A, Fortuno C, Gómez-Sanz A, Varga MJ, Chamberlin A, Richardson ME, Michailidou K, Pérez-Segura P, Spurdle AB, de la Hoya M. ACMG/AMP interpretation of BRCA1 missense variants: Structure-informed scores add evidence strength granularity to the PP3/BP4 computational evidence. Am J Hum Genet. 2025;112(5):993– 1002. doi:10.1016/j.ajhg.2024.12.011

